# Direction-specific excitation-inhibition imbalances and their neurobiological substrates in recurrent major depressive disorder

**DOI:** 10.64898/2026.05.17.26351714

**Authors:** Sihan Ding, Huaijin Gao, Rui Qian, Baorong Gu, Dan Wu, Zhiyong Zhao

## Abstract

**Background:** Major depressive disorder (MDD) is characterized by disrupted information flow among brain regions. While effective connectivity (EC) captures these causal interactions, the underlying structural and molecular basis remain unclear. This study aims to investigate direction-specific EC alterations in MDD and their associations with laminar structural covariance (SC) and transcriptional and neurotransmitter profiles.

**Methods:** Resting-state fMRI and structural MRI data were analyzed from the REST-meta-MDD consortium (Discovery, N=1627) and an independent cohort (Validation, N=226). We calculated the unsigned and signed EC using Liang Information Flow and laminar SC based on cortical depth, and compared them between MDD patients and healthy controls. The EC alterations were further associated with molecular profiles integrating gene expression (AHBA) and neurotransmitter receptors (PET/SPECT). Then, Chain mediation analyses were performed to map the hierarchical pathways from molecular basis to EC. Finally, we evaluated the clinical potential of EC in its therapeutic responses to medication and neuromodulation in a longitudinal dataset (N = 16 for medication, N = 11 for neuromodulation).

**Results:** Our analysis revealed no significant changes in the EC of first-episode MDD but observed a hyper-driven cerebellar-cerebral EC pattern in recurrent MDD (RMDD), characterized by a direction-specific excitation-inhibition imbalance featuring enhanced inhibitory cerebellar output alongside a concurrent increase in both inhibitory input and excitatory output within sensorimotor/cognitive regions. These alterations were physically constrained by specific laminar SC patterns, particularly involving the middle cortical lamina. Moreover, the input EC changes in RMDD patients were primarily enriched in biological processes related to the modulation of chemical synaptic transmission, whereas output EC changes were linked to synapse structure regulation. These EC alterations were closely associated with serotonergic, GABAergic, and glutamatergic neurotransmitter systems. Importantly, we identified oligodendrocyte precursor cells (OPCs) as a key cellular mediator bridging microscale molecular features to macroscale connectional alterations in RMDD. These findings were reproducible in the validation dataset. Clinically, medication treatment primarily evoked a pattern of decreased input coupled with increased output, whereas neuromodulation elicited a reciprocal shift characterized by enhanced input and attenuated output.

**Conclusions:** These findings underscore a direction-specific gene-neurotransmitter-cell type-laminar SC-EC pathological model in RMDD. By integrating multi-scale biological mechanisms with clinical phenotypes, this study highlights the potential of directional EC as a biomarker for stratifying refractory depression and guiding precision therapeutics.

## 1. Introduction

Major depressive disorder (MDD) is a severe psychiatric illness characterized by widespread and complex patterns of brain dysfunction, imposing a substantial socioeconomic and health burden globally.^1^ Notably, MDD exhibits significant clinical and neurobiological heterogeneity, particularly between first-episode drug-naïve (FEDN) and RMDD patients. Compared to those in their first episode, patients with RMDD often experience more severe somatic symptoms, greater impairments in cognitive functions and mental representation processing, and an increased risk of suicidal ideation.^2^ These clinical disparities are mirrored by distinct neurophysiological profiles, as evidenced by recent research identifying divergent spatiotemporal trajectories and connectivity patterns across different MDD subtypes.^3,4^ Despite these well-documented differences in both clinical presentation and neurophysiological markers, the underlying neurobiological mechanisms that distinguish these subtypes remain poorly understood. To explore these neural substrates, resting-state functional MRI (rs-fMRI) has been extensively utilized, with previous studies predominantly focusing on functional connectivity (FC), characterized by disrupted FC across large-scale networks, specifically manifesting as hyperconnectivity within the default mode network (DMN) and hypoconnectivity within the frontoparietal network (FN) in MDD.^5^ RMDD is uniquely characterized by complex dysconnections including long-range hyperconnectivity across sensorimotor and cognitive regions, which are often absent in FEDN patients.^2^ The patients with first-episode MDD typically manifest more extensive and intensive FC reductions across the DMN, cognitive control network, and affective network compared to RMDD patients.^6^ Additionally, FEDN cases have been shown to exhibit a specific increase in FC between the amygdala and right prefrontal cortex, which significantly decreases and tends to normalize following clinical remission.^7^

However, due to the inherently undirected nature of traditional FC, it fails to characterize the causal interactions between brain regions.^8^ In contrast, effective connectivity (EC) captures the directionality and causal influence of neural signaling, enabling a more precise characterization of pathological information flow patterns.^8,9^ Previous neuroimaging studies have reported disrupted EC patterns in MDD, indicating altered directional communication across large-scale brain networks^10^. Specifically, investigations into the sensorimotor hierarchy have demonstrated that depression is associated with significantly altered top-down (backward) and bottom-up (forward) signaling.^11^ Subtype-targeted research further underscores the mechanistic superiority of EC, revealing that RMDD involves a “hyper-driven” efferent (output) flow from the vlPFC alongside reduced cerebellar input, whereas FEDN patients primarily exhibit increased directional communication from the sensorimotor network (SMN) to the fronto-parietal network.^12^ By capturing the causal influence and directionality of neural signaling, EC provides enhanced sensitivity for clinical stratification, enabling machine learning models to achieve diagnostic accuracies of up to 94.35%, which significantly outperforms the 60-70% range typically reported for FC-based models.^13^ However, while these macroscopic pathological information flow patterns are increasingly well-mapped, the underlying laminar structural and molecular mechanisms remain largely elusive.

The link between brain structural architecture and functional activity is well-established at the macroscale, where anatomical organization provides the physical substrate that constrains neural signaling, while functional activity can, in turn, reshape these structures via neuroplasticity.^14,15^ Prior neuroimaging studies have largely overlooked these relationships at the cortical laminar level, often treating the cortex as a single homogenous unit. However, recent research has increasingly underscored the critical importance of fine-scale cortical organization. Histological evidence demonstrates that inter-regional patterns of laminar structure—such as laminar thickness covariance—are closely associated with the strength of functional connections.^16^ Meanwhile, recent high-resolution neuroimaging research has further established that the FC is highly sensitive to the fine-grained laminar connectional architecture of the human brain.^17–21^ For instance, within the visual system, rs-fMRI has identified specific “output-to-input” layer connections, such as those from V1 Layer II/III to MT Layer IV, and feedforward projections from superficial-intermediate V1 to intermediate-deep V2.^20^ Beyond these specialized circuits, large-scale resting-state networks also exhibit distinct depth-specificity; for example, communication within the DMN is primarily supported by intermediate and superficial layers.^19^ Furthermore, recent multilayer network analyses have revealed that superficial depths dominate whole-brain information transfer, whereas deeper layers drive network clustering.^18^ These findings collectively indicate that the distinct tangential layers of the isocortex are viewed as fundamental units of complex information processing, which support hierarchical message passing and cortical organization through specific feedforward and feedback projection patterns.^16,17,20^. However, there remains a critical gap in our understanding of how fine-scale laminar structural configurations support or constrain the directionality of information flow. Furthermore, the direct associations between laminar-specific structural substrates and the directional EC alterations characterizing MDD has yet to be fully elucidated.

While MDD is characterized by widespread abnormalities in the structure, function, and coordinated activity of large-scale brain networks^22^, these macroscopic neuroimaging phenotypes are fundamentally governed and shaped by underlying molecular and cellular processes.^23^ Consequently, unraveling the microscale substrates that drive these macroscale circuit dysfunctions is imperative for both advancing our understanding of MDD pathophysiology and identifying targets for precision-psychiatry interventions. In recent years, the emergence of comprehensive brain-wide gene expression atlases (e.g., the Allen Human Brain Atlas, AHBA^24^) and positron emission tomography (PET)-derived neurotransmitter receptor maps^25^ has provided an unprecedented capacity to directly link spatially resolved molecular features—including gene expression profiles, receptor densities, and cell-type specificity—to in vivo neuroimaging markers, thereby bridging the gap between molecular-scale variations and systems-level organization.^25,26^ Previous research has established a robust molecular foundation for MDD. Large-scale genome-wide association studies (GWAS) have identified hundreds of risk variants enriched in biological processes such as nervous system development, synapse assembly, and synaptic function.^27–29^ At the neurochemical level, MDD involves complex dysregulation across serotonergic, dopaminergic, glutamatergic, and GABAergic systems.^22,30,31^ Cell-specific investigations have implicated somatostatin (SST) interneurons, astrocytes, and OPCs as pivotal biological substrates that modulate circuit stability and mediate depressive phenotypes.^23,32^ Recent evidence further suggests that FEDN is characterized by a localized enrichment of genes related to synaptic plasticity and potentiation in the prefrontal cortex,^33^ recurrent and chronic illness is associated with more pervasive neurochemical deficits, such as a more widespread downregulation of mGluR5.^31,34^ Despite these insights linking molecular profiles to brain structure and functional, the specific molecular mechanisms driving the directional specificity of EC alterations, especially whether information inflow and outflow dysfunctions stem from distinct genetic and neurochemical substrates, remain unclear.

Accordingly, this study aims to investigate direction-specific EC alterations among MDD subgroups and to explore their potential laminar-specific structural and molecular substrates using multimodal neuroimaging (Figure 1). Using cross-sectional resting-state fMRI and T1-weighted imaging data from the REST-meta-MDD cohort (discovery dataset), we first calculated EC via Liang Information Flow as well as laminar SC based on KL divergence. We then evaluated subtype differences in demographics, EC, SC and the associations between EC alterations and genetic, cell-specific transcriptomic as well as neurotransmitter profiles. After that, we conducted laminar-level correlation and chain mediation analyses, bridging microscale structural and molecular pathologies with macroscopic circuit dysfunctions. Finally, we evaluated the clinical utility of EC by validating their treatment response to medication and neuromodulation in a longitudinal dataset. We hypothesized that: (1) FEDN and RMDD patients exhibit divergent pathological information flow patterns, specifically characterized by a “hyper-driven” cerebellar-cerebral EC in RMDD; (2) these alterations are physically constrained by fine-grained laminar structural configurations rather than simple one-to-one mappings; (3) the directionality of EC alterations (input vs. output) is governed by distinct genetic, cellular, and neurochemical substrates; and (4) can act as sensitive biomarkers for monitoring treatment responses to both medication and neuromodulation, providing a novel framework for precision diagnosis and stratified treatment strategies.

**Figure 1:**
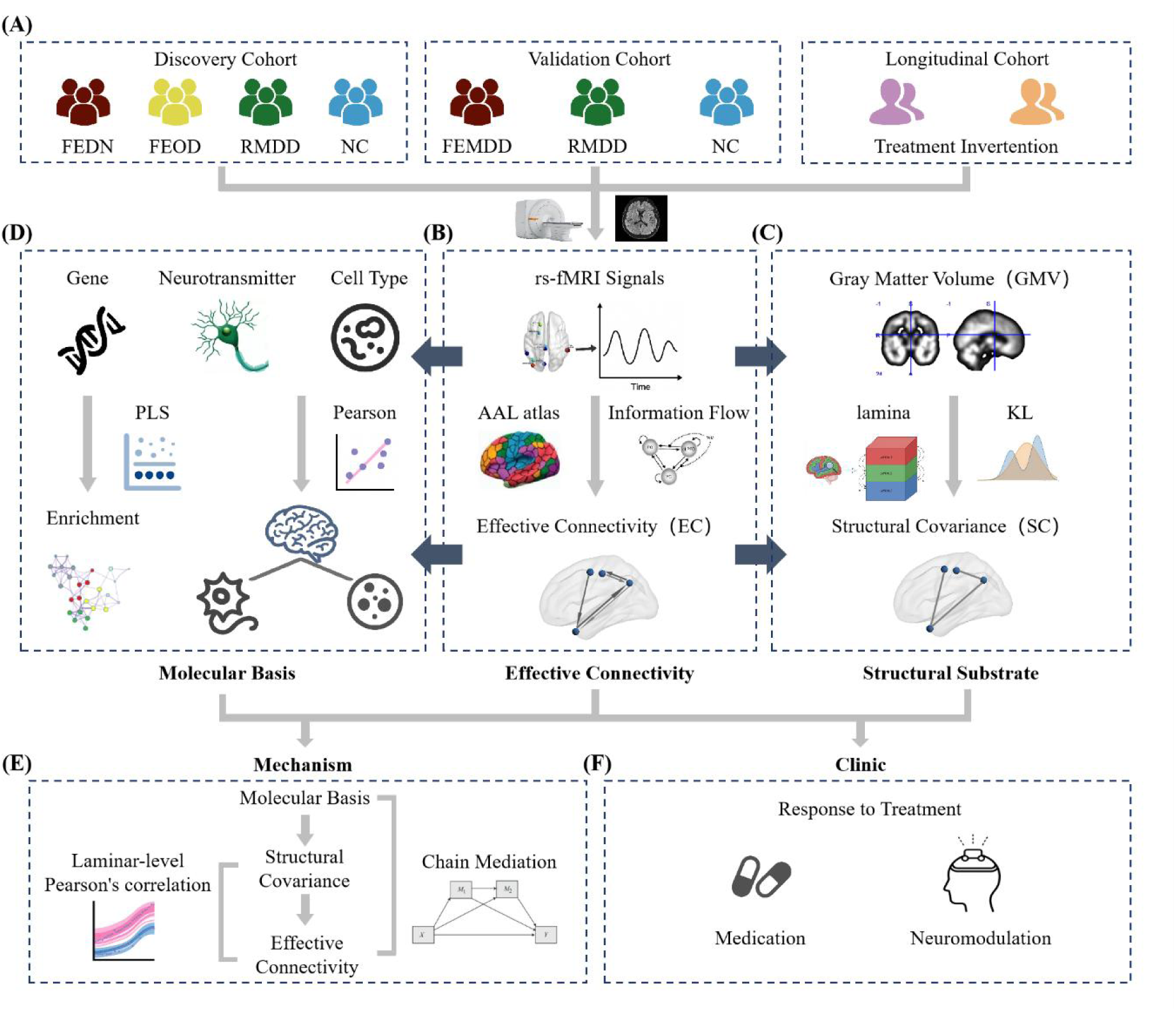
Workflow of the study. **(A) Cohort Characterization**: The study included a discovery cohort from the REST-meta-MDD project, alongside an independent validation cohort and a longitudinal cohort. **(B) EC Analysis**: Direction-specific connectivity alterations were identified from resting-state fMRI data using the Liang Information Flow method. **(C) Structural Subtrate**: Laminar SC was calculated based on T1-weighted GMV. **(D) Molecular Basis**: Biological substrates of EC alterations were investigated by integrating multimodal molecular profiles, including neurotransmitter receptor densities (PET/SPECT) and transcriptomic data (AHBA). **(E) Multiscale Mechanism**: Correlation and chain mediation analyses were employed to delineate the pathways spanning from the molecular basis to laminar structural organization and macro-scale EC alterations. **(F) Clinical Significance**: The utility of EC features was evaluated for predicting longitudinal responses to medication and neuromodulation.

## 2. Methods

### 2.1 Data acquisition

In the discovery cohort, strict inclusion and exclusion criteria were applied. Subjects were required to be aged 18–65 with complete demographic information. We excluded sites containing late-onset or remitted patients, as well as individual subjects with inadequate brain coverage, excessive head motion (mean FD > 0.5 mm), or poor spatial normalization quality. Sites with fewer than 10 subjects per group were also removed to ensure statistical robustness. Finally, resting-state fMRI and T1-weighted images were obtained from the large-sample REST-meta-MDD dataset^35^, including five paired subgroup comparisons: total MDD patients vs. Normal Controls (NC) (n = 839 vs. 788), First-Episode Drug-Naïve (FEDN) patients vs. NCs (n = 227 vs. 388), Recurrent MDD (RMDD) patients vs. NCs (n = 189 vs. 423), FEDN patients vs. RMDD patients (n = 117 vs. 72), and FEDN patients vs. First-Episode On-Drug (FEOD) patients (n = 227 vs. 100). All study sites obtained approval from their local institutional review boards and ethics committees, and all participants provided written informed consent.

The validation cohort included 86 First-Episode MDD (FEMDD), 33 RMDD and 84 NCs from the First Affiliated Hospital of Zhejiang University. Notably, although the validation cohort included Chinese participants, it was independent of all sites listed in the REST-meta-MDD Consortium. To further assess the treatment response of EC, we collected longitudinal fMRI data of 27 MDD patients, which were categorized into the Medication Treatment Group (n = 16) and the Neuromodulation Group (n = 11) based on the intervention received. Detailed protocols for the treatments were provided in the Supplementary Materials. Clinical scale scores, including HAMD, HAMA, BSI5, BSI14, BSI19, and SHAPS, were collected before and after treatment.

### 2.2 Genetic, Neurotransmitters, and Cellular profiles

Gene transcriptional data were obtained from the AHBA (http://human.brain-map.org)^24^. The Abagen toolbox (https://github.com/rmarkello/abagen) was employed to map the gene expression data onto 116 brain regions of automated anatomical labeling (AAL) atlas^36^. Following standard pipelines, microarray probes were re-annotated, low-quality probes were removed, and expression values were normalized across samples. For each donor brain, samples were assigned to the corresponding cortical parcels, and expression values were averaged across donors to yield a 116 region × 15,633 gene matrix.

The spatial distributions of 18 neurotransmitter receptors and transporters were obtained from a public PET dataset, including serotonergic (5-HT1a, 5HT1b, 5-HT2a, 5-HT4), cannabinoid (CB1, CBF), dopaminergic (D1, D2, DAT, F-DOPA), GABAergic (GABAa), opioid (kappaOp, MU), noradrenergic (NAT), glutamatergic (NMDA, mGluR5), serotonergic transporter (SERT), and cholinergic (VAChT) systems.

To estimate the cellular landscape across the brain, we utilized transcriptomic data from the AHBA to quantify the densities of six principal cell types: neurons, astrocytes, oligodendrocytes, microglia, endothelial cells, and oligodendrocyte precursor cells (OPCs). Specifically, we applied a feature gene decomposition-based deconvolution method via the BRETIGEA software package (https://github.com/andymckenzie/BRETIGEA), leveraging canonical genetic markers to calculate cell-type proportions^37–39^. These molecular profiles were then mapped onto the 116 brain regions defined by the AAL atlas, providing a comprehensive whole-brain reference of cellular abundance.

### 2.3 EC calculation

All fMRl and structural T1-weighted MRI images were preprocessed consistently at each site followed a standard pipeline including: slice-timing correction, realignment, DARTEL-based MNI normalization, nuisance regression, spatial smoothing (6 mm FWHM), and temporal bandpass filtering (0.01–0.1 Hz). Detailed procedures were summarized in the literature^35^.

The EC, reflecting the information outflow and inflow intensity, was calculated based on 116 regions of AAL atlas^36^ using Liang Information Flow^40^, which rigorously derived from first principles and quantifies the causality between two time series by measuring the transfer rate of information.

EC was quantified using Liang information flow, a rigorous causal inference method derived from the first principles of information entropy and dynamical systems theory.^40^ Unlike empirical measures such as Granger causality, this approach offers significant computational advantages by relying solely on analytical formulas involving sample covariances and temporal derivatives.^40–43^ Furthermore, the method is model-free and does not require the assumption of stationarity, which is particularly valuable for rs-fMRI data where the underlying interaction structures are often unknown or non-stationary.^40,41^ Crucially, the Liang method provides a signed measure of information transfer: positive causality indicates that the source makes the target more uncertain (excitatory or facilitative effect), while negative causality signifies that the source makes the target more certain (inhibitory or suppressive effect).^40^ This unique property allows for a more fine-grained understanding of the excitation-inhibition (E/I) balance within MDD neural circuits. This framework has demonstrated remarkable robustness in extreme conditions, such as systems buried in heavy noise or networks of nearly synchronized chaotic oscillators, where traditional methods often fail to differentiate confounding processes.^42^ By capturing these hierarchical and directional interactions, recent research has achieved high classification accuracy in identifying complex neurodevelopmental disorders, underscoring the method’s potential for precision diagnosis.^40^

Specifically, for two normalized time series *X* _1_ and *X* _2_, the rate of information flow from *X* _2_ to *X* _1_ (*T* _2 → 1_) is calculated as follows^40^:

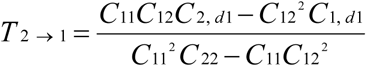

where *C*_ij_ represents the sample covariance between *X*_i_ and *X*_j_, and *C_i_*_, *d*_*_j_* is the covariance between *X*_i_ and the coupled differential signal 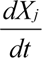. A positive *T* _2 → 1_ indicates an excitatory effect where the source increases the uncertainty of the target, while a negative value signifies an inhibitory regulation that makes the target more certain. Notably, the regional EC was defined as the average intensity of a specific brain region to the whole brain in input or output information flow.

To exploit the signed property of Liang Information Flow, where positive and negative values represent excitatory and inhibitory effects respectively, we decomposed connection-level EC into four direction- and sign-specific ROI-level metrics. Specifically, for each of the 116 AAL regions, we calculated the average excitatory input (input_pos), inhibitory input (input_neg), excitatory output (output_pos), and inhibitory output (output_neg). For inhibitory metrics (negative values), the absolute values were used to represent the magnitude of inhibitory intensity for subsequent statistical comparisons. This procedure yielded a 116×4 metric matrix per participant, enabling a fine-grained characterization of regional information transfer profiles.

### 2.4 Laminar SC construction

The boundaries of GM-CSF (pial surface) and GM-WM (WM surface) were obtained from T1-weighted images by dilating the GM and WM masks by one voxel using FSL (https://fsl.fmrib.ox.ac.uk/fsl/fslwiki/) and intersecting the dilated images with the original GM mask. The pial surface, WM surface, and GM mask were then input into the LAYNII tools^44^ to divide the entire GM cortex into three equidistant laminae (superficial, middle and deep) for each case. The innermost and outermost laminae were excluded to reduce segmentation errors due to partial volume effects. Laminar structural covariance (SC) was then constructed by calculating the Kullback-Leibler divergence between gray matter volume (GMV) probability density functions (PDF)^45^ across regions and laminae. Specifically, the statistical similarity between two PDFs (p and q) was quantified using a symmetric version of KL divergence (*KL*(*p*, *q*)):

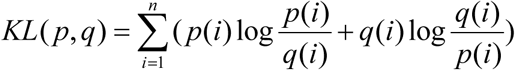

The resulting divergence was then transformed into a similarity measurement (KLS) using an exponential function / the formula *KLS* (*p*, *q*) = *e*^−^*^KL^*^(^ *^p^*^,*q*)^, ensuring a value range between 0 and 1.

### 2.5 Statistical analysis

To visualize the normative EC distribution in healthy controls, we averaged the ROI-level positive and negative EC strengths across NC subjects, separately for input and output directions. This yielded four EC maps: positive input, positive output, negative input, and negative output, which were used to characterize the baseline spatial pattern of excitatory and inhibitory information flow in the NC group.

Group differences in EC and SC were compared between paired groups using Linear Mixed-Effect (LME) model, with site as a random factor and age, sex, education, and head motion as covariates. Associations between EC and laminar SC were assessed using Pearson’s correlations. Multiple comparisons were controlled using false discovery rate (FDR) correction (p < 0.05). We also performed LME to evaluate the associations between EC and clinical symptom severity, as measured by HAMD and HAMA scores.

Partial Least Squares (PLS) regression was performed to associate regional EC alterations (response variables) with gene expression profiles (predictor variables). We employed spatial autocorrelation-corrected permutation testing (1000 iterations) to assess the statistical significance of the primary component (PLS1). To identify the most influential contributors to PLS1, bootstrapping (5000 iterations) was used to estimate the standard error and calculate Z-scores for each gene. The top 1000 genes ranked by |Z| were then subjected to functional enrichment analysis using Metascape, covering GO-BP, GO-CC, and KEGG pathways. Terms surviving FDR correction (p < 0.01) were considered significant and were retained for interpretation. Spatial correlations between altered EC and neurotransmitter density or cell type were examined using Juspace^46^ with Pearson’s correlation coefficients across 116 regions. Statistical significance was assessed using permutation tests (1000 permutations) with p < 0.05 considered significant.

Chain mediation analysis was performed to explore the hierarchical pathways linking molecular profiles to imaging phenotype alterations, using the PROCESS function in the bruceR R package (https://CRAN.R-project.org/package=bruceR). Genes with the top three positive and negative weights for each direction were incorporated into the model, along with significant neurotransmitter and cell types. After standardizing all variables with Z-score transformation, a multi-step chain model was conducted to test the sequential indirect effects from gene expression (X) to EC (Y), through intermediate mediators including neurotransmitter density (M1), cell-type distributions (M2) (or vice versa), and SC (M3). Statistical significance of the indirect effects was determined via non-parametric bootstrapping with 1000 iterations. A chain mediation effect (Path: *X* → *M*_1_ → *M* _2_ → *M* _3_ → *Y*) was considered significant if the 95% bias-corrected confidence interval (CI) did not encompass zero.

### 2.6 Treatment response analysis

To assess clinical improvement following intervention, paired-sample t-tests were performed to compare clinical scale scores between pre-treatment and post-treatment time points. Subsequently, the therapeutic responses of EC to medication and neuromodulation were evaluated using two-way repeated-measures analyses of covariance (ANCOVA). For these analyses, regional EC was categorized into three distinct metrics: unsigned EC, positive EC (comprising excitatory input and output), and negative EC (calculated using the absolute values of inhibitory input and output to represent magnitude). Each of the three metrics was subjected to a separate time (pre-treatment vs. post-treatment) × direction (Input vs. Output) interaction model. In all models, EC served as the dependent variable while controlling for age, gender, and education as covariates. Multiple comparisons were corrected using the FDR method with a threshold of p < 0.05. For significant time × direction interactions, post hoc paired-sample t-tests were conducted within each direction (Input and Output) to identify specific longitudinal changes in neural signaling.

### 2.7 Validation analysis

To evaluate the robustness and generalizability of our main findings, we conducted the following additional validations: (1) EC and laminar SC analyses in an independent validation dataset; (2) the spatial correlations between EC alterations and gene expression, neurotransmitter densities, and cell-type distributions by replacing the AAL atlas with the CC200 atlas^47^.

## 3. Results

### 3.1 Demographic and Clinical Characteristics

In the discovery cohort, total MDD patients and NCs were matched in age (p > 0.05). However, MDD patients had significantly shorter years of education compared to NCs (p < 0.001) and a higher proportion of females (p = 0.005). Both FEDN and RMDD patients had shorter years of education than NCs (all p < 0.001). Additionally, while FEDN patients exhibited a shorter duration of illness (p < 0.001) than RMDD patients, there were no significant differences between them in age, gender, education, and HAMD scores. The validation sample included 119 MDD patients (86 FEDN and 33 RMDD) and 84 NCs. Both FEDN and RMDD groups showed higher score in HAMD and HAMA, and shorter years in education compared to NC (p < 0.001). Three groups had no significant differences in age and gender.

### 3.2 EC differences between groups

The NC group-averaged EC maps (Figure 2A) revealed distinct spatial distributions of regional input and output strengths for both positive and negative EC components, highlighting the direction- and sign-dependent organization of effective information flow in the healthy brain. Compared with NCs, the RMDD group showed increased EC inputs to the precentral gyrus, postcentral gyrus, angular gyrus and middle temporal gyrus (MTG), as well as increased EC outputs from the cerebellum (Figure 2B). A significant EC difference in the MTG was also observed between the FEDN and FEOD groups, suggesting a potential medication-related effect. The sign-specific analysis provided deeper insights into the previously identified unsigned EC abnormalities. Specifically, for the sensorimotor and cognitive hubs, the decomposition clarified that these were driven by a significant diminution of both inhibitory input (input_neg) and excitatory output (output_pos). Similarly, the EC output disruptions in the cerebellum were clarified to be primarily driven by a weakened inhibitory output (output_neg), suggesting a significant loss of cerebellar-mediated suppressive control over the cerebral cortex in RMDD. More details are provided in Supplementary Material. No significant EC differences were observed in the other group comparisons. No significant correlations were found between EC and clinical symptom severity (HAMD or HAMA scores) across any of the patient groups.

**Figure 2:**
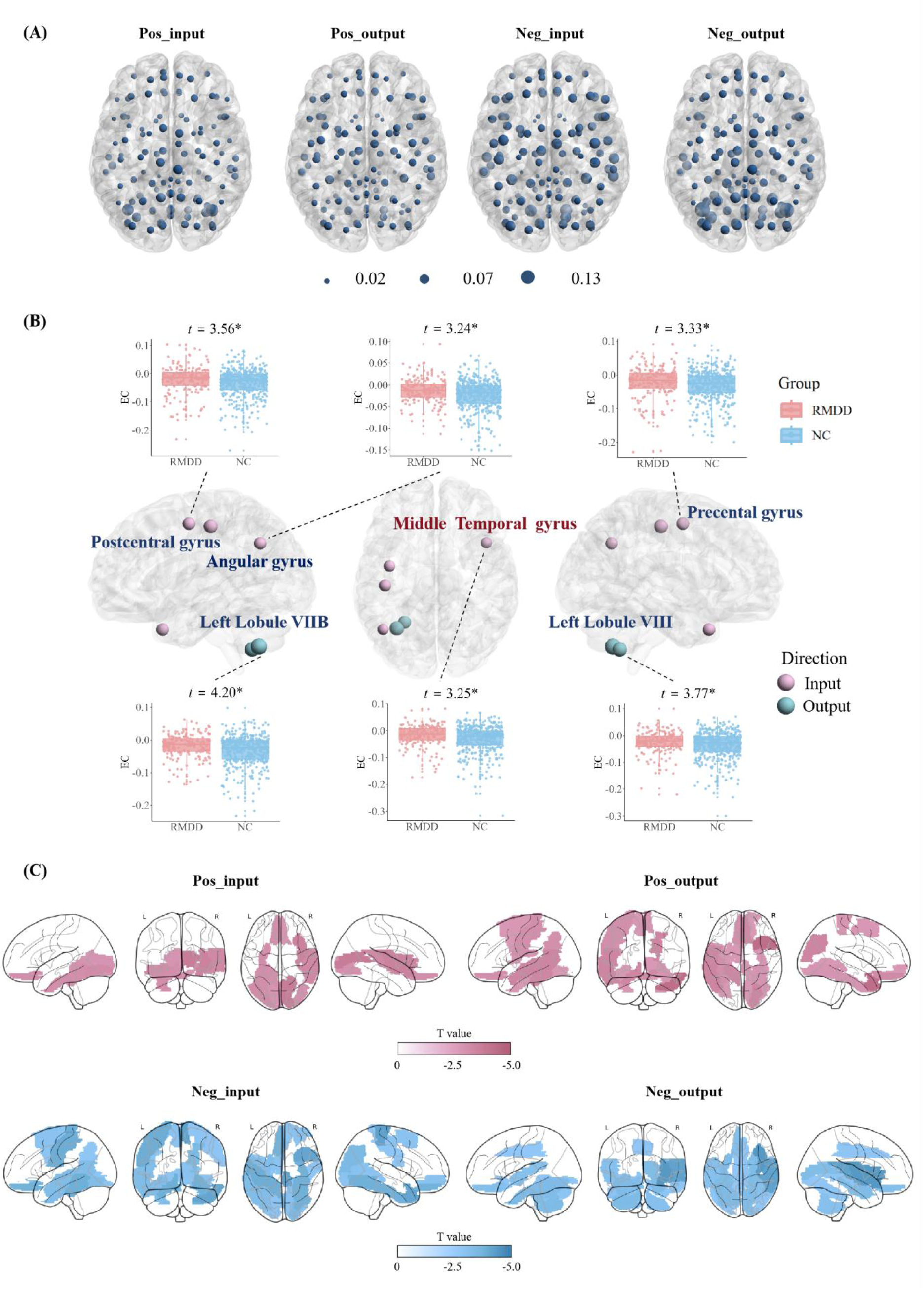
EC differences between groups. (A) NC group-averaged ROI-level EC maps showing positive and negative EC strengths in the input and output directions. The node size represents the raw regional EC intensity. **(B) EC differences at the regional level.** Regions that survived after FDR correction are shown, along with each t-value derived from LME. The middle temporal gyrus is labeled with red font, as it also differs between FEOD and FEDN. **(C) Sign-EC difference.** Regions that survived after FDR correction are shown. *: 0.01 ≤ adjusted-*p* < 0.05.

LME models identified widespread regional alterations across all four metrics (Figure 2C) as follows: (1) Excitatory Input (Pos_input): Alterations were localized within 11 regions primarily associated with the visual-affective integration system, including the orbital part of the frontal cortex, visual processing areas, and cerebellar lobule VI; (2) Excitatory Output (Pos_output): 16 regions demonstrated significant differences, concentrated in areas governing primary perception and spatial coordination, such as the paracentral lobule, sensorimotor cortex, and temporal regions. (3) Inhibitory Input (Neg_input): Significant changes were identified in 19 regions across the motor-sensory-visual system, encompassing the precentral and postcentral gyri, the angular gyrus, and multiple temporal and visual hubs; (4) Inhibitory Output (Neg_output): This metric exhibited the most extensive alterations within 22 regions, primarily involving high-level language, auditory, and social semantic hubs, alongside a dense cluster in the cerebellar system, including the superior temporal gyrus and temporal pole. Substantial spatial overlaps were observed between the significant regions of input_pos and output_neg, as well as between input_neg and output_pos. Notably, several core pathological hubs exhibited significant alterations across all four indicators simultaneously, including the rectal gyrus, fusiform gyrus, and the left cerebellar lobule VI.

### 3.3 The SC differences and Correlations between EC and laminar SC

We observed lower SC between the middle lamina of middle cingulum gyrus and the middle lamina of fusiform gyrus, and higher SC between the middle lamina of superior parietal lobule and the middle lamina of superior temporal gyrus in the RMDD group compared with NCs (Figure 3A). No significant SC differences were observed in the other group comparisons.

**Figure 3:**
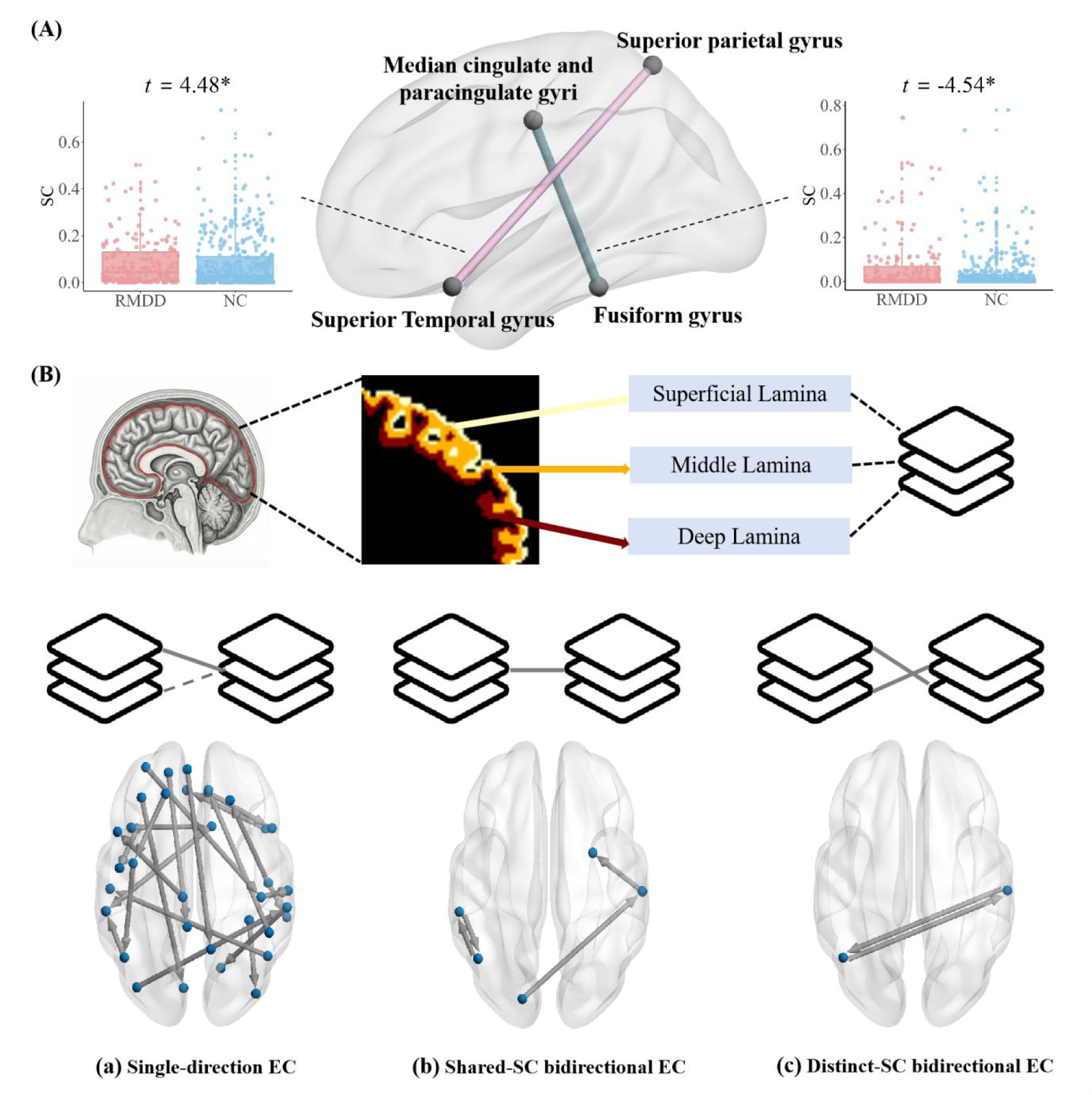
SC differences and Associations between EC and laminar SC. (A) SC differences at the laminar level. Connections that survived after FDR correction are shown, along with each t-value derived from LME. **(B) Associations between EC and laminar SC.** Regions and connections survived after FDR correction are shown. For bidirectional EC between any pair of brain regions, three distinct patterns of structural support are identified: (a) Single-direction EC showed significant associations with only one direction of EC but not the other; (b) Shared-SC bidirectional EC were commonly correlated with bidirectional ECs, such as connections between occipital and temporal lobes and between the supramarginal and angular gyri; (c) Distinct-SC bidirectional EC showed different associations with bidirectional ECs, such as the temporal-angular connection. *: 0.01 ≤ adjusted-*p* < 0.05.

The relationships between EC and laminar SC showed three distinct patterns (Figure 3B): (a) Single-direction EC, where only one EC direction within a regional pair showed a significant relationship with laminar SC; (b) Shared-SC bidirectional EC, where bidirectional ECs were both associated with the same laminar SC, including occipital–temporal and supramarginal–angular connections; (c) Distinct-SC bidirectional EC, where the two directions of EC were associated with different laminar SCs, such as the temporal–angular connection.

### 3.4 Associations between EC alterations and genes, neurotransmitter and cell type, and chain mediation analysis

The first component of PLSR (PLS1) in both input and output directions captured the dominant gene–EC associations differentiating RMDD from NCs. In the input direction, PLS1 explained 35.2% of the variance (p < 0.05), whereas in the output direction it explained 29.3% (p = 0.055). In both cases, the regional PLS1 scores were significantly positively correlated with the corresponding EC T-maps (input: r = 0.593; output: r = 0.542; both p < 0.001, permutation tests with spatial autocorrelation correction). Gene-wise loadings revealed direction-specific molecular profiles. Input-related EC alterations were associated with genes such as TENM2 and CFAP65 (positive) and CLCN5 and GALNT13 (negative), with GO enrichment highlighting processes related to the **modulation of chemical synaptic transmission** (Figure 4A). In contrast, output-related EC alterations were linked to genes including IL11 and CLEC11A (positive) and ZWINT (negative), and were enriched for **synapse structure regulation and cell junction organization** (Figure 4B).

**Figure 4:**
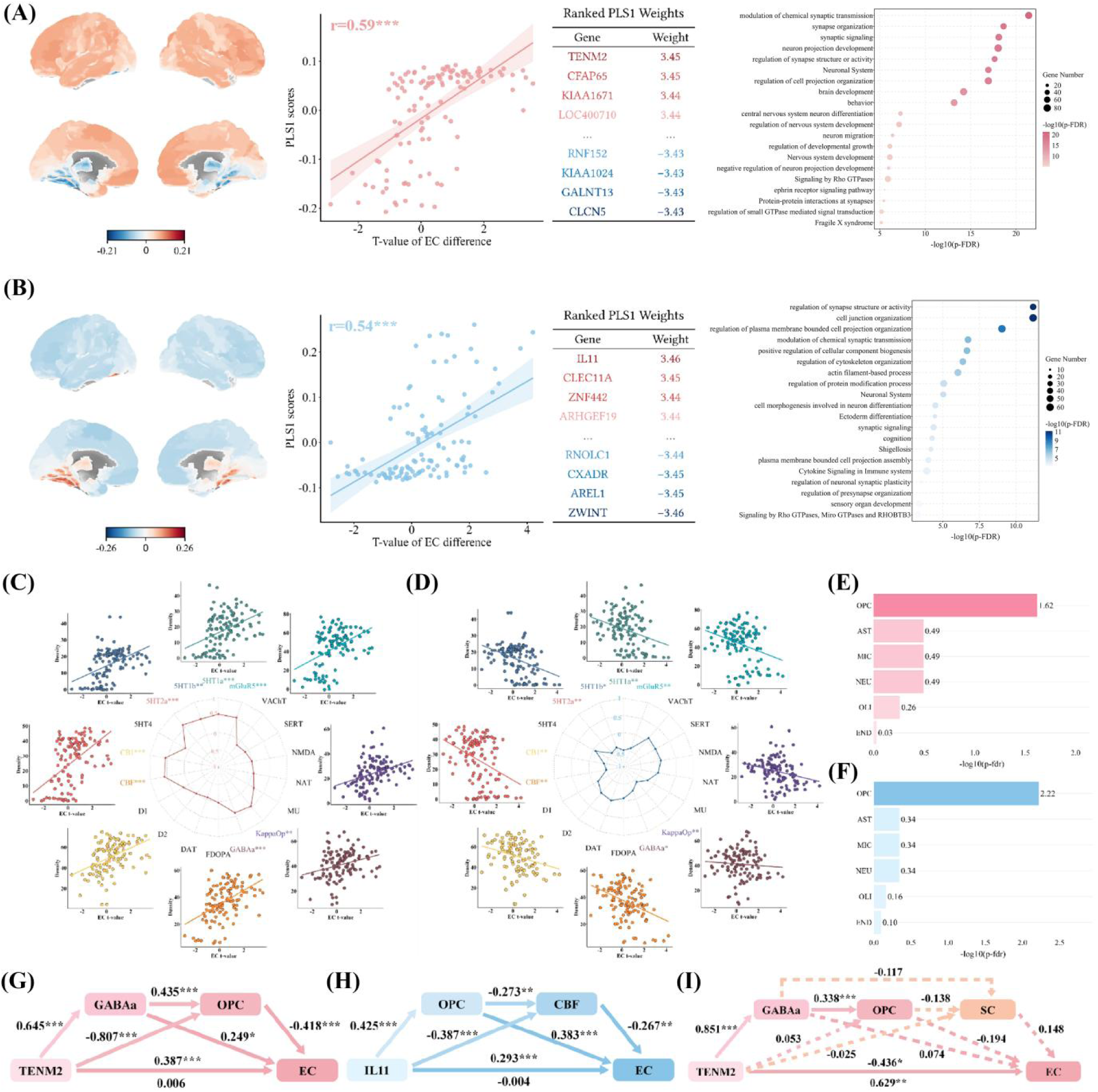
Associations and chain mediation model examples between EC alterations and molecular profiles in RMDD compared with NC. (A) Associations between gene expression and EC alterations in input and output direction (B). The PLS1 score maps for each direction are shown using the AAL atlas, with corresponding scatter plots provided. The positively and negatively weighted PLS1 values of genes are shown in the table, along with the enriched gene terms and pathways for altered EC. (C) Spatial correlations between EC alterations in input and output direction (D) and neurotransmitter densities across brain regions. The colorful labels except black represent significant permutation-based p-values after FDR correction, while radar charts and scatter plots show the Pearson correlation coefficient. (E) Spatial correlations between EC alterations in input and output direction (F) and cell types across brain regions. (G) Input and Output (H) direction without SC as M3. (I) Input direction with SC as M3. *: 0.01 ≤ adjusted-p < 0.05; **: 0.001 ≤ adjusted-p < 0.01; ***: adjusted-p < 0.001.

For the FEDN vs. NC comparison (Supplementary Figure S3B), the PLS1 for input-related EC explained 18.9% of the variance (p=0.044). In the output direction, the sixth component (PLS6) explained the most variance (11.0%), though this association was not statistically significant (p=0.175). Enrichment analysis for the input direction in FEDN patients showed substantial overlap with the RMDD group, specifically highlighting biological processes related to the modulation of chemical synaptic transmission and the Neuronal System. In contrast, enrichment pathways for output-related EC in the FEDN group did not survive correction for multiple comparisons.

While the spatial correlations of EC-neurotransmitter density exhibited **opposite signs** between input and output directions—all correlations in the input direction were positive, whereas those in the output direction were negative— the involved neurotransmitter categories of EC alterations in RMDD remained highly consistent, including serotonergic (5-HT1a, |r|=0.51-0.53; 5-HT1b, |r|=0.39-0.48; 5-HT2a, |r|=0.46-0.59), cannabinoid (CB1, |r|=0.43-0.55; CBF, |r|=0.50-0.58), GABAergic (GABA-a, |r|=0.26-0.39), glutamatergic (mGluR5, |r|=0.43-0.52), and opioid (KappaOp, |r|=0.43-0.46) markers (all adjusted-p < 0.05; Figure 4C, 4D). In contrast, FEDN displayed a largely overlapping neurotransmitter profile for input EC, whereas its output EC alterations exhibited a differentiated signature with significant correlations linked to noradrenergic (NAT) and glutamatergic (NMDA) systems but lacked associations with the CBF and mGluR5 observed in RMDD (Supplementary Figure S3C).

At the cellular level, EC differences between RMDD and HC were selectively associated with the spatial distribution of oligodendrocyte precursor cells (OPCs) in both input (r = -0.450, adjusted-p = 0.024) and output (r = 0.463, adjusted-p = 0.006) directions (Figure 4E, 4F). This pattern was also identified in FEDN vs. NC (Supplementary Figure S3D).

For path of Gene→neurotransmitter→cell→EC, we identified 10 chained models in the output direction and 6 of the output using the T-values derived from the RMDD vs. NC comparison. For path of Gene→cell→neurotransmitter→EC, we identified 16 chained models in the output direction and 19 of the output using the T-values derived from the RMDD vs. NC comparison. In contrast, no significant chain mediation effects were identified for the FEDN vs. NC comparison. The example chain mediation models for the RMDD group are shown in Figure 4. In the input direction, TENM2 significantly positively correlated with the GABAa (β = 0.645, p < 0.001), which in turn significantly positively correlated with OPC (β = 0.435, p < 0.001), and consequently, significantly negatively correlated with EC (β = -0.418, p < 0.001). Critically, bootstrapped analyses revealed that the indirect effect (i.e. TENM2→GABAa→OPC→EC) was significant (Figure 4G). In the output direction, IL11 significantly positively correlated with the OPC (β=0.425, p<0.001), which in turn significantly negatively correlated with CBF (β=-0.273, p<0.01), and consequently, significantly negatively correlated with EC (β=-0.267, p<0.01). Critically, bootstrapped analyses revealed that the indirect effect (i.e. IL11→OPC→CBF→EC) was significant (Figure 4H). However, when considering SC as the mediator M3, no serial model show a significant indirect effect (Figure 4I). All other identified models were listed in Table X (Supplementary material).

### 3.5 EC response to medication and neuromodulation treatment

Both interventions showed significant clinical improvements. Patients showed a marked alleviation of core affective distress after medication treatment, with decreases in both depression (HAMD: p < 0.001) and anxiety (HAMA: p < 0.001) levels, although such changes were observed in suicidal ideation metrics or anhedonia (Figure 5A). Meanwhile, the neuromodulation group demonstrated a more extensive pattern of improvement (Figure 5B). Beyond the substantial reductions in depression (HAMD: p < 0.001) and anxiety (HAMA: p < 0.001), patients also achieved significant improvements suicidal ideation (BSI-5, 14, and 19) and anhedonia (SHAPS: p < 0.001).

**Figure 5:**
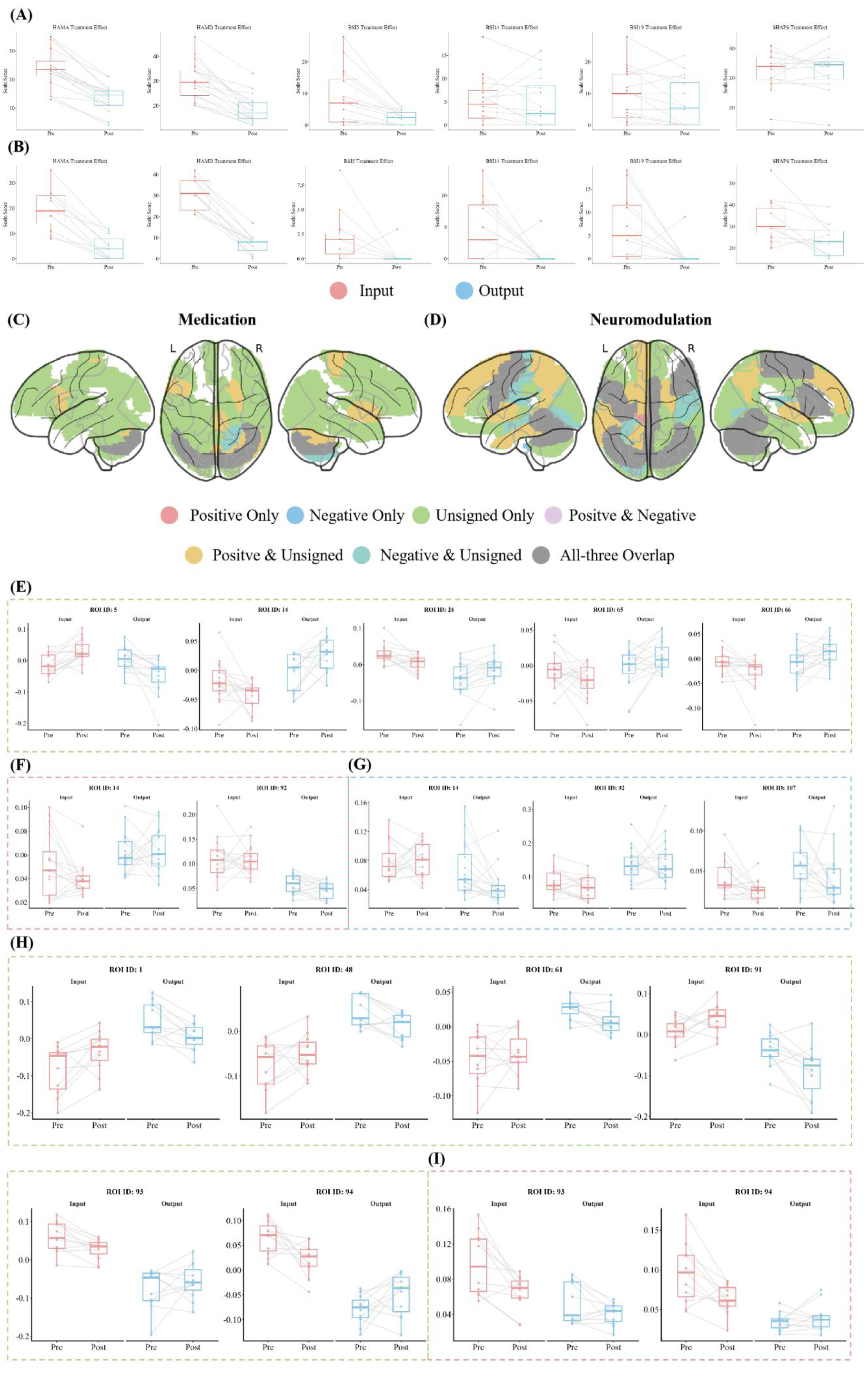
Response to medication and neuromodulation treatment. (A) Clinical scale scores alterations after treatment using medication and (B) neuromodulation. (C) Brain regions showing significant time × direction interaction effects of unsigned, positive, and negative EC associated with response to escitalopram monotherapy and (D) neuromodulation, respectively (adjusted-p < 0.05). (E) Specific significant alterations within each brain regions for unsigned EC, (F) positive EC, (G) and negative EC in the Medication Group. (H) Specific significant alterations within each brain regions for unsigned EC and (I) positive EC in the Neuromodulation Group.

The repeated-measures ANCOVA of EC revealed widespread significant time × direction interaction effects for both treatment after FDR correction (p<0.05). For the medication group, significant interactions were identified in 60 regions for unsigned EC, 21 regions for positive EC, and 20 regions for negative EC, with varying degrees of spatial overlap among the three metrics (Figure 5C). For the neuromodulation group, significant interactions were observed in 50 regions for unsigned EC, 7 regions for positive EC, and 3 regions for negative EC (Figure 5D). Post hoc paired-sample t-tests further clarified that for the medication group, in the unsigned EC analysis, both input and output directions showed significant changes in the prefrontal cortex and angular gyrus. When considering the sign of EC, the positive EC showed significant changes in the input of the right triangular part of inferior frontal gyrus and the output of the right cerebellum, while the negative EC showed such changes in the input of the bilateral cerebellum and the output of the right triangular part of inferior frontal gyrus. As for the neuromodulation group, in the unsigned EC analysis, significant input changes were identified in the left precentral gyrus and bilateral cerebellum, while more output changes were observed in the left precentral gyrus, right lingual gyrus, left inferior parietal gyrus, and left cerebellum. For positive EC, significant changes were found in the input of bilateral cerebellum, with no significant findings in the output. No significant changes were identified for the negative EC in the neuromodulation group. Collectively, these interaction patterns may suggest direction- and sign-dependent differences in the neural response to distinct therapies. Specific alterations within each brain region were shown in Supplementary Figure S4.

### 3.6 Validation results

The independent validation dataset revealed increased EC input to the MTG and output from the cerebellum, which aligns with the main discovery. Laminar SC support for EC presented three distinct patterns same as the discovery (Supplementary Figure S1), further demonstrating it varies across brain regions. Associations between EC alterations and molecular substrates remained stable using the CC200 atlas (Supplementary Figure S2).

## 4 Discussion

In this study, we provide a comprehensive, multi-scale characterization of the directional information flow disruptions and their underlying biological substrates across different subtypes of MDD. In contrast to the absence of significant EC alterations in FEDN, RMDD exhibited a pronounced “hyper-driven” cerebellar-cerebral circuitry characterized by increased causal inputs to sensorimotor and cognitive hubs alongside augmented outputs from the cerebellum. Further sign-specific decomposition revealed that these abnormalities reflected a disruption of excitation-inhibition balance, with substantial overlaps between altered excitatory and inhibitory directional signaling, suggesting impaired feedback-dependent regulation within large-scale neural circuits.

Importantly, longitudinal analyses further demonstrated that directional EC is sensitive to therapeutic intervention. Medication and neuromodulation induced distinct patterns of EC reorganization, indicating different forms of direction-dependent neuroplastic recovery.

These EC abonormalities are physically constrained by fine-grained laminar SC architectures—particularly involving the middle cortical lamina—through unidirectional, shared, and distinct structural-functional association patterns. At the microscale, these directional dysfunctions were associated with distinct molecular signatures: input-related anomalies are primarily enriched in chemical synaptic transmission pathways, whereas output-related disruptions are linked to synapse structural regulation, with GABAergic, serotonergic and cannabinoid systems, as well as OPCs serving as pivotal mediators bridging these scales. Collectively, our findings support a cross-scale framework linking directional EC dysfunction and treatment-related EC reorganization to their structural and molecular substrates in RMDD.

### 4.1 Direction-specific excitation-inhibition imbalances in MDD subtypes

Our results revealed altered information flows predominantly in RMDD rather than FEDN, suggesting that RMDD may involve more stable and widespread directional dysconnectivity than the FEDN stage. This subtype-specific pattern is consistent with previous REST-meta-MDD evidence^12^ suggesting that early-stage depression may involve relatively circumscribed sensorimotor-to-control network reweighting, while RMDD may be accompanied by broader cerebellar-cortical and cognitive-control reorganization. The increased ECs input to the sensorimotor regions we observed were consistent with recent evidence^48^, suggesting that the sensorimotor integration is a core physiological signature of depression, potentially underlying the embodied symptomatology of the disorder. Meanwhile, the augmented EC outputs from the cerebellum support the “hyper-driven cerebellar-cerebral” circuit model.^49^ Recent multi-site EC-based classification work further identified cerebellar and fronto-temporal EC features as highly discriminative for MDD and emphasized that many key features originated from cerebellar regions^50^. Together with prior brain-wide EC findings^51^ implicating temporal, orbitofrontal, hippocampal, precentral, and postcentral regions in MDD, our altered signaling to temporal and angular hubs may reflect impaired integration of social, semantic, and self-related information, as well as disrupted homeostatic communication required for goal-directed executive control.

By leveraging the signed properties of Liang Information Flow, we further characterized RMDD as a disorder of sign- and direction-specific information flow rather than a simple abnormality of overall EC magnitude. In signed EC frameworks, positive and negative directed coefficients reflect whether one region predicts an increase or decrease in another region’s subsequent activity, providing a macroscopic description of excitatory-like and inhibitory-like causal influence between brain regions^52^. This interpretation is consistent with previous MDD studies using Granger causality or DCM, which reported altered positive/negative causal influences across limbic, prefrontal, sensorimotor, and high-order networks^53^.

Notably, the substantial overlap between input_pos and output_neg, as well as between input_neg and output_pos, suggests that RMDD does not affect excitatory-like or inhibitory-like information flow independently. Instead, the same regions may show paired abnormalities across opposite signed directions, indicating a disruption of feedback-like regulation. In a balanced system, excitatory-like input may need to be counteracted by inhibitory-like output, while inhibitory-like input may be accompanied by compensatory excitatory-like output. Therefore, the observed overlap may reflect impaired negative-feedback coordination between incoming and outgoing signed information flow. This interpretation is supported by prior resting-state DCM evidence showing that MDD involves sign- and direction-specific alterations in large-scale EC rather than a uniform increase or decrease in communication strength^54,55^.

Several regions, including the rectal gyrus, fusiform gyrus, and cerebellar lobule VI, showed significant alterations across all four signed EC indicators. This convergence suggests a more severe local disturbance of E/I-like information-flow balance, in which both input and output, positive and negative components are jointly affected. Similar signed EC abnormalities have been reported in depression-related sensorimotor hierarchies, default-mode and salience networks, and emotional face-processing circuits, where altered forward/backward or excitatory/inhibitory causal influences were associated with symptom severity, treatment response, or impaired top-down regulation^11,54,56,57^. Together, these findings suggest that RMDD may involve a breakdown of local signed-flow homeostasis in key perceptual, affective, and regulatory hubs.

### 4.2 Direction- and sign-dependent EC responses to clinical interventions

Our longitudinal findings showed that both medication and neuromodulation improved clinical symptoms, but they induced distinct EC reconfiguration patterns. Medication was mainly characterized by decreased input and increased output, whereas neuromodulation showed enhanced input and attenuated output. Beyond this direction-dependent pattern, signed decomposition further revealed intervention-specific responses: medication affected both positive and negative EC, whereas neuromodulation mainly involved unsigned and positive EC changes, with limited negative-EC alterations.

These findings highlight the added value of signed EC. Direction-dependent EC analysis captures whether treatment mainly changes afferent or efferent information flow, whereas sign-dependent analysis further distinguishes whether these changes are driven by positive or negative information transfer. This distinction is important because previous EC studies indicate that depression-related treatment effects are not merely changes in overall connectivity strength, but involve forward and backward EC abnormalities in sensorimotor hierarchies that can partially revert toward group-average levels after treatment, suggesting that EC can track clinical course and treatment response^11^. Similarly, treatment-related changes in brain connectivity have been repeatedly reported across antidepressant, ECT, and rTMS studies, with clinical improvement associated with connectivity changes in most reviewed studies^58^.

The sign-dependent findings provide a more refined interpretation of the two interventions. The medication group showed changes in both positive and negative EC, suggesting a broader recalibration of facilitative and suppressive information flow. This is compatible with prior evidence that antidepressant treatment can alter cortical regulatory systems, including increased occipital GABA after SSRI treatment in MDD and downstream GABA/glutamate-related effects after escitalopram administration^59,60^. These neurochemical findings support the biological plausibility that pharmacological treatment may influence both positive and negative components of brain communication.

By contrast, neuromodulation primarily affected unsigned and positive EC, suggesting a more selective reshaping of excitatory-like information transfer and input-output balance. This interpretation is consistent with TMS studies showing that clinical response is associated with changes in distributed connectivity pathways rather than only local stimulation-site effects. A recent systematic review of MRI connectivity predictors of TMS response in MDD emphasized that treatment effects involve the sgACC, executive-control, salience, and default-mode networks, with marked heterogeneity across studies^61^. MRS studies further suggest that TMS can affect E/I-related neurobiology, including increased medial prefrontal GABA after left DLPFC rTMS, while sham-controlled iTBS evidence indicates that such effects may be region- and individual-dependent rather than uniform^62,63^. This may explain why neuromodulation in our cohort showed prominent positive-EC changes but limited negative-EC changes.

Together, these findings suggest that signed EC provides a more sensitive framework for characterizing treatment-related neural plasticity in MDD than unsigned EC alone. Medication may induce broader recalibration across both positive and negative information flow, whereas neuromodulation may preferentially reshape positive and direction-specific communication within sensorimotor – cerebellar circuits. Thus, the combination of direction and sign offers a more precise biomarker for distinguishing intervention-specific EC responses.

### 4.3 Structural and molecular basis of EC directional dysfunction in RMDD

We found the reduced SC between the middle lamina of the middle cingulum and fusiform gyrus in RMDD, and no significant SC alterations in FEDN. Although this specific pathway has rarely been described, prior MDD-SCN studies have convergently reported disrupted covariance across cingulate-, temporal-, DMN-, salience-, and prefrontal-centered networks^64–66^. Our finding therefore extends these macro-scale SCN abnormalities to the laminar level, suggesting a recurrent-stage vulnerability of cingulate-temporal/visual structural coordination. Crucially, we identified three distinct association patterns between directional EC and laminar SC, which may support the framework that altered information flow in RMDD may be constrained by lamina-dependent structural connection, rather than a simple one-to-one mapping^16,17^. This interpretation is consistent with recent evidence showing that laminar thickness covariance is related to both EC-based cortical hierarchy and interregional structural and functional connectivity^16^. The Unidirectional and Distinct SC patterns may reflect the asymmetric nature of hierarchical communication, as feedforward and feedback projections have distinct laminar profiles and are associated with separable electrophysiological signatures^67,68^. The Shared SC pattern, in contrast, may be related to the “similar prefers similar” wiring principle, whereby regions with similar laminar or cytoarchitectonic profiles tend to show stronger connectivity^16,69^. Together, these findings suggest that laminar SC provides a region- and direction-sensitive structural scaffold for EC abnormalities in RMDD.

Gene expression analysis revealed distinct molecular substrates underlying directional EC alterations in RMDD. Input-related EC abnormalities were enriched in pathways related to chemical synaptic transmission, whereas output-related alterations were associated with synapse structure regulation and cell junction organization, consistent with the view that MDD is fundamentally a synaptic disorder involving disrupted synaptic transmission and plasticity^27^. Among the key genes, TENM2 showed the strongest positive association in the input direction and is known to organize inhibitory synapses and recruit GABAA receptors via EB protein-mediated microtubule interactions^70^. In contrast, IL11 emerged as a key output-related gene and has been implicated in OPC trophic support and neuroimmune regulation within the CNS^71^. These findings suggest that distinct genetic programs may contribute to the directional specificity of EC dysfunction in RMDD. Our EC-neurotransmitter association analysis found that EC bidirectional alterations were both correlated to serotonergic (5-HT1a, 5-HT1b, 5-HT2a), GABAergic (GABAa) and glutamatergic (mGluR5) systems. Specifically, 5-HT1a regulates serotonergic firing and stress-related neuroendocrine responses^72–74^, whereas 5-HT2a modulates cortical excitatory activity and emotional processing^75^. The association with GABAa receptors supports impaired inhibitory regulation in depression^76^, while mGluR5 is closely linked to synaptic plasticity and stress resilience^34^. Together, these neurotransmitter systems may provide the neurochemical basis through which molecular vulnerabilities translate into the macroscopic EC disruptions characterizing RMDD. At the cellular level, OPC distribution showed the strongest spatial association with EC alterations, consistent with previous studies^77^. OPCs uniquely receive direct glutamatergic and GABAergic synaptic inputs and express receptors such as GABAa and mGluR5^78,79^. In addition, OPCs actively participate in circuit remodeling through synaptic and axonal engulfment^80^. The spatial correlation implies that EC disruptions in RMDD may stem from a maladaptive interplay at the neuron-glia synapse. These observations suggest that, despite distinct neuroimaging and clinical features, input and output directions may share common cellular and neurochemical mechanisms contributing to their EC alterations, underscoring the complex, multi-level pathophysiology of MDD.

Our chain mediation analyses revealed distinct biological pathways bridging micro-scale molecular abnormalities to macro-scale EC dysfunction, supporting a multi-level cascade model of MDD pathophysiology. In the input direction, the significant TENM2→GABAa→OPC→EC pathway suggests that TENM2-related deficits may disrupt inhibitory synaptic organization and GABAa receptor localization^70^, thereby impairing OPC regulation through altered GABAergic signaling^78,79^ and ultimately contributing to EC abnormalities^81^. In the output direction, the IL11→OPC→CBF→EC pathway indicates that IL-11-dependent OPC maintenance^71^ may influence endocannabinoid signaling, potentially through lipid metabolic processes critical for cannabinoid system function^32^. Given the role of endocannabinoids in retrograde synaptic regulation and long-term depression^82^, this pathway may represent a mechanism buffering against maladaptive connectivity patterns in depression. Interestingly, significant mediation effects were primarily observed in pathways without SC involvement, whereas models incorporating SC did not survive significance testing. This finding suggests that EC abnormalities in MDD may more directly reflect rapid synaptic and neurochemical dysfunctions driven by genes, neurotransmitters, and cell types, rather than slower large-scale structural covariance changes.

### 4.4 Limitations and future directions

Several limitations in the present study should be acknowledged. First, the sample size for the longitudinal analysis was relatively small, which may limit the generalizability of treatment-related findings. Future studies should include larger cohorts to better capture individual variability in treatment response. Second, although we validated the stability of the neurophysiological subtypes using an external dataset, the validation sample was limited in size. Further replication in larger independent datasets, particularly those encompassing diverse populations in terms of ethnicity, age, and clinical presentation, is essential to establish the robustness and broader applicability of our findings. Third, research on EC and its laminar SC and molecular basis in depression remains limited. Future studies should replicate our findings and examine how our EC approach relates to conventional estimation methods, such as DCM and GCM. Fourth, detailed medication data were unavailable for patients in the REST-meta-MDD dataset, which limited our ability to fully control for potential confounding effects of antidepressant use. Future work should investigate how different classes, dosages, and durations of antidepressant treatment may influence EC alterations. Finally, the neuroimaging, transcriptomic, and neurotransmitter data used in this study were not obtained from the same individuals. This non-matched, integrative approach may introduce potential biases and complicate the interpretation of cross-modal associations. Future studies incorporating multimodal data from the same participants will be crucial for more precise characterization of the biological underpinnings of depression subtypes.

## 5 Conclusion

This study is based on the multi-center REST-meta-MDD dataset and the independent validation set, and systematically reveals the direction-specific EC disorder and its cross-scale biological mechanisms in recurrent depression (RMDD). Our results indicate that RMDD presents an abnormality in the over-driven cerebellar-brain directional connection, and this macroscopic functional reorganization does not occur randomly but is subject to the physical constraints of the laminar SC, showing a hierarchical-specific structural-function support pattern. At the molecular level, multimodal molecular analysis reveals the heterogeneous pathological pathways underlying the different direction EC abnormalities. Clinical analysis further confirms that this direction-dependent EC feature reflects the different neuroplasticity response patterns of medication treatment and neural regulation. In summary, this study integrates genes, neurotransmitters, cell types, laminar SC, and EC to propose a cross-scale MDD pathological model, revealing the neurobiological characteristics of severe depression, and providing a new perspective for developing biomarker-based precision diagnosis and stratified treatment strategies.

## Data Availability

Data of the REST-meta-MDD project are available at : http://rfmri.org/REST-meta-MDD. Neurotransmitter receptor and transporter data can be obtained online at https://github.com/juryxy/JuSpace/tree/JuSpace_v1.5/JuSpace_v1.5/PETatlas. Human gene expression data that support the findings of this study are available in the Allen Brain Atlas (https://human.brain-map.org/static/download).

